# Cost-Outcome Variation in Percutaneous Mechanical Circulatory Support: A National Value-of-Care Analysis

**DOI:** 10.64898/2026.08.31.26361851

**Authors:** Joshua D. Greendyk, William E. Allen, Afif Hossain, Jonathan R. Lopez, Zachariya Trichas

## Abstract

**Background:** Percutaneous mechanical circulatory support (pMCS) is increasingly used in critically ill patients, yet its value in relation to cost and outcomes remains unclear. We evaluated national variation in utilization, outcomes, and cost, and introduced a value-of-care framework integrating risk-adjusted outcomes and expenditures.

**Methods:** We performed a retrospective cohort study using the National Inpatient Sample to identify non-elective hospitalizations of critically ill patients undergoing intra-aortic balloon pump (IABP) or percutaneous left ventricular assist device (pLVAD) placement using ICD-10 codes. Multivariable logistic regression and generalized linear models were used to estimate expected outcomes and costs. Observed-to-expected (O/E) ratios were calculated, and a value index was derived to compare procedural strategies.

**Results:** A total of 57,910 weighted hospitalizations were included (IABP 78%, pLVAD 22%). In-hospital mortality exceeded 30% across regions. Significant regional variation was observed, with the West demonstrating the highest costs and the Midwest the lowest (p<0.001). Mean hospital charges were higher for pLVAD compared with IABP ($403,731 vs $320,769). Both strategies achieved outcomes better than expected after risk adjustment (O/E 0.92); however, costs were higher than expected for both, with greater relative cost inflation observed for IABP (O/E 1.41) and higher absolute costs for pLVAD. In value-of-care analysis, IABP was associated with lower cost and comparable outcomes, while pLVAD demonstrated higher cost without proportional outcome improvement.

**Conclusion:** Substantial variation exists in the cost, outcomes, and value of pMCS strategies. While both IABP and pLVAD achieve favorable risk-adjusted outcomes, pLVAD is associated with higher costs without commensurate clinical benefit.

## Introduction

Healthcare delivery in the United States is increasingly challenged by persistent concerns surrounding outcomes, quality, patient safety and cost. Cardiovascular disease and stroke (CVDS) remain the leading causes of morbidity and mortality in the United States and are major contributors to national healthcare expenditures, accounting for approximately $251 billion in spending in 2019 (1). This financial burden carries significant downstream consequences, as medical expenses account for more than half of personal bankruptcies in the United States, including among insured individuals (2). Despite this disproportionate investment, higher healthcare spending has not consistently translated into improved clinical outcomes or enhanced quality of care, underscoring a critical disconnect between cost and value within the current system (3).

This challenge is particularly evident in the management of critically ill patients, where resource-intensive interventions are frequently employed (4). Percutaneous mechanical circulatory support (pMCS) devices have become increasingly utilized in patients with cardiogenic shock and hemodynamic instability (5). Contemporary strategies include the Impella, which provides direct left ventricular unloading; the TandemHeart, which enables left atrial to arterial bypass with higher levels of circulatory support; and the Intra-aortic balloon pump, a historically established modality with more modest hemodynamic augmentation. The intra-aortic balloon pump (IABP) is generally used for temporary, lower-level hemodynamic support and coronary perfusion augmentation in cardiogenic shock or peri-procedural ischemia, whereas the Impella provides more robust left ventricular unloading and cardiac output support for severe cardiogenic shock or high-risk PCI (6). TandemHeart is typically reserved for profound refractory cardiogenic shock requiring higher-flow circulatory support via left atrial-to-arterial bypass (6). While these devices offer varying degrees of support, their use is associated with substantial costs, prolonged intensive care utilization, and a significant risk of complications, contributing to wide variability in both outcomes and resource utilization (5, 6).

Despite the rapid expansion of pMCS use, existing literature is largely limited to single-center experiences or registry-based analyses that primarily focus on clinical outcomes without integrating economic considerations. As a result, there remains a critical gap in understanding how these interventions perform from a value perspective, particularly at a national level.

Evaluating both outcomes and costs in a risk-adjusted framework is essential to inform clinical decision-making and optimize resource allocation. Accordingly, this study leverages the National Inpatient Sample (NIS) to perform a cost-outcome analysis of patients undergoing pMCS using standardized ICD-10 procedural coding. We introduce a novel “care value” framework that integrates risk-adjusted outcomes with expected and observed costs to assess the relative value of these interventions. By comparing device strategies within a unified cost-outcomes model, this study aims to identify meaningful variation in value across pMCS modalities and provide a foundation for more efficient, value-based cardiovascular care.

## Methods

### Cohort selection

We conducted a retrospective cohort study using the National Inpatient Sample (NIS), the largest all-payer inpatient database in the United States, which provides nationally representative estimates of hospitalizations. Patients were identified using ICD-10-PCS codes for emergent percutaneous hemodynamic support, including insertion of percutaneous left ventricular assist devices (pLVAD) (02HA3RZ) and physiologic circulatory support codes (5A02110, 5A0221D), capturing devices such as Impella and TandemHeart, as well as intra-aortic balloon pump placement (5A02210). Patients were grouped into a IABP and pLVAD cohorts to compare demographics by procedure. Cost analysis included the total cost for the hospital stay. Further analyses stratified by hospital region to capture regional differences. We included adult patients undergoing these procedures during non-elective admissions with evidence of critical illness (Figure 1). Critical illness was defined by the presence of cardiogenic shock, cardiac arrest, or receipt of life-support interventions including mechanical ventilation or mechanical circulatory support. To focus on early, index interventions, procedures were restricted to those performed on hospital day 0 or 1 when timing data were available. Patients receiving multiple percutaneous hemodynamic interventions during the same hospitalization were excluded. All analyses incorporated discharge-level sampling weights provided by the NIS to generate nationally representative estimates.

**Figure 1:**
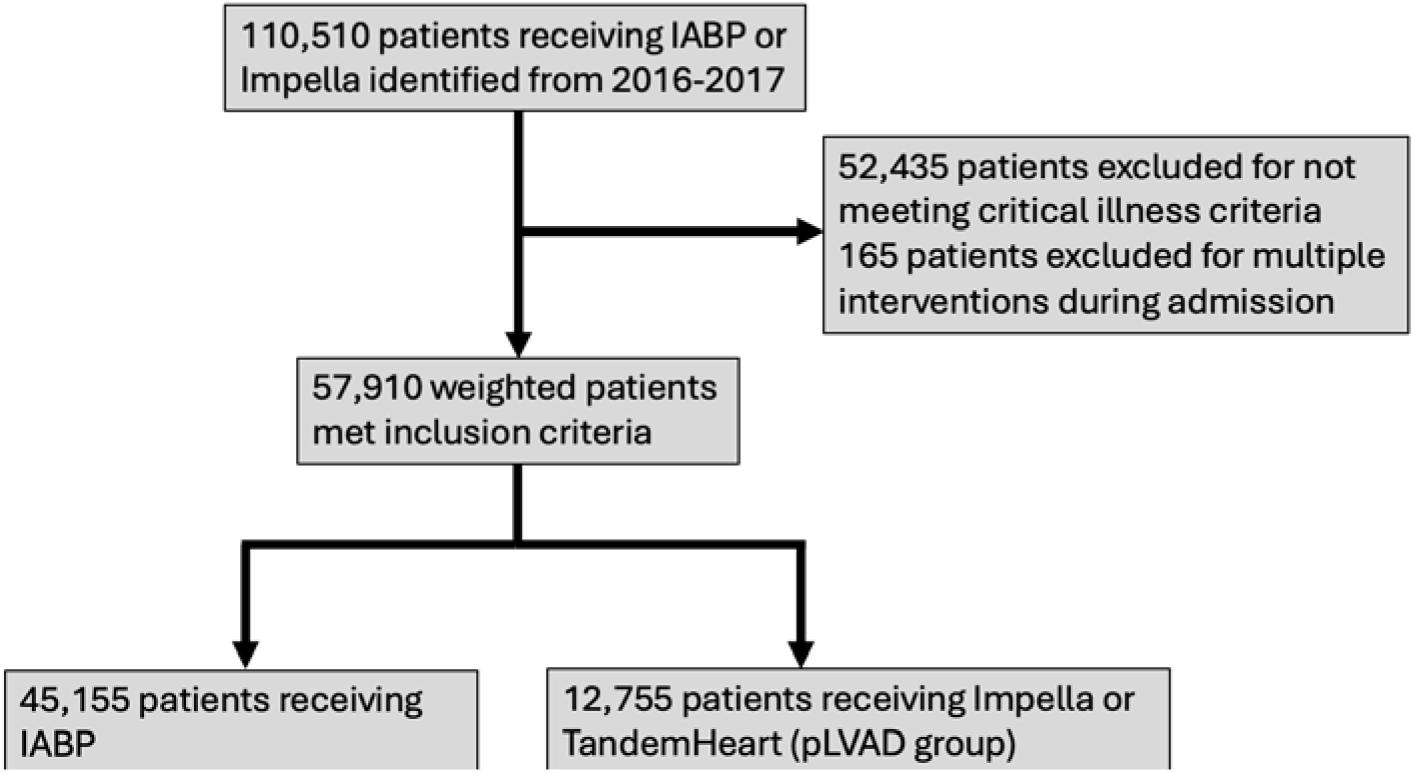
Patient selection

### Value of care analysis

Value of care was assessed by jointly evaluating risk-adjusted hospital charges and clinical outcomes across procedure types. Expected hospital charges were estimated using a generalized linear model with a gamma distribution and log link to account for right-skewed cost data. Expected risk of adverse clinical outcomes was estimated using multivariable logistic regression. Observed-to-expected (O/E) ratios were calculated for both cost and outcomes at the procedure level. A value index was derived to quantify relative cost burden in relation to outcome burden, enabling comparison of value across procedural strategies.

### Statistical analysis

Cohort demographics stratified by procedure type or hospital region were evaluated using both univariate and multivariable analyses. Univariate comparisons were performed using Pearson chi-square tests for categorical variables and analysis of variance (ANOVA) for continuous variables. Multivariable regression models were used to assess the association between hospital characteristics and both costs and clinical outcomes, with results reported as adjusted estimates, odds ratios (ORs), and corresponding confidence intervals (CIs). All models adjusted for patient- and hospital-level covariates, including demographics, comorbidities, and hospital characteristics. Logistic regression was used for binary outcomes, and generalized linear models were used to estimate adjusted costs. Results are presented with appropriate measures of uncertainty. All statistical analyses were performed using IBM SPSS Statistics version 25.0 (IBM Corp., Armonk, NY), and a two-sided p-value <0.05 was considered statistically significant.

## Results

A total of 57,910 weighted hospitalizations met inclusion criteria, including 45,155 (78.0%) patients undergoing intra-aortic balloon pump (IABP) placement and 12,755 (22.0%) receiving percutaneous left ventricular assist devices (pLVAD) (Figure 1). Patients in the pLVAD cohort were slightly younger (64.0 vs 65.3 years, p<0.001) and more frequently male (70.9% vs 69.1%, p<0.001). Racial distribution was similar between groups, with most patients identifying as White (72.6% vs 70.7%), followed by Black (11.0% vs 11.1%) and Hispanic (8.7% vs 9.5%) populations. Comorbidity burden differed modestly between cohorts, with higher rates of congestive heart failure in the pLVAD group (64.1% vs 59.2%), while hypertension (69.5% vs 64.0%), diabetes (40.6% vs 38.1%), atrial fibrillation (35.1% vs 31.1%), and valvular disease (23.9% vs 18.9%) were more prevalent among IABP patients (all p<0.001). Both cohorts were predominantly treated at urban teaching hospitals (77% overall) and large centers (>63%), with regional variation noted, including greater pLVAD utilization in the South (42.0%) and West (21.8%) compared with the Midwest (19.4%) and Northeast (16.7%) (p<0.001) (Table 1).

**Table 1:**
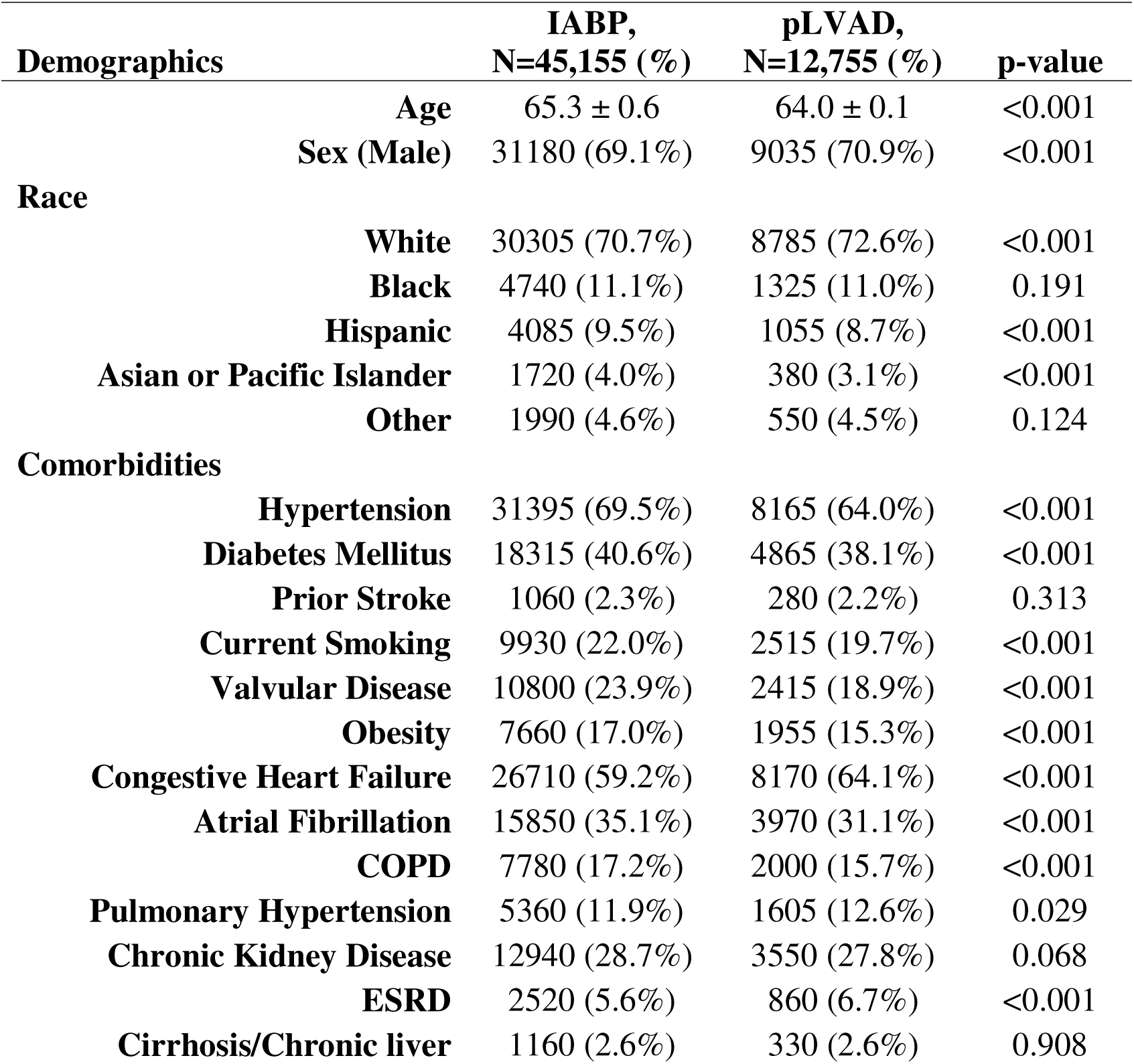
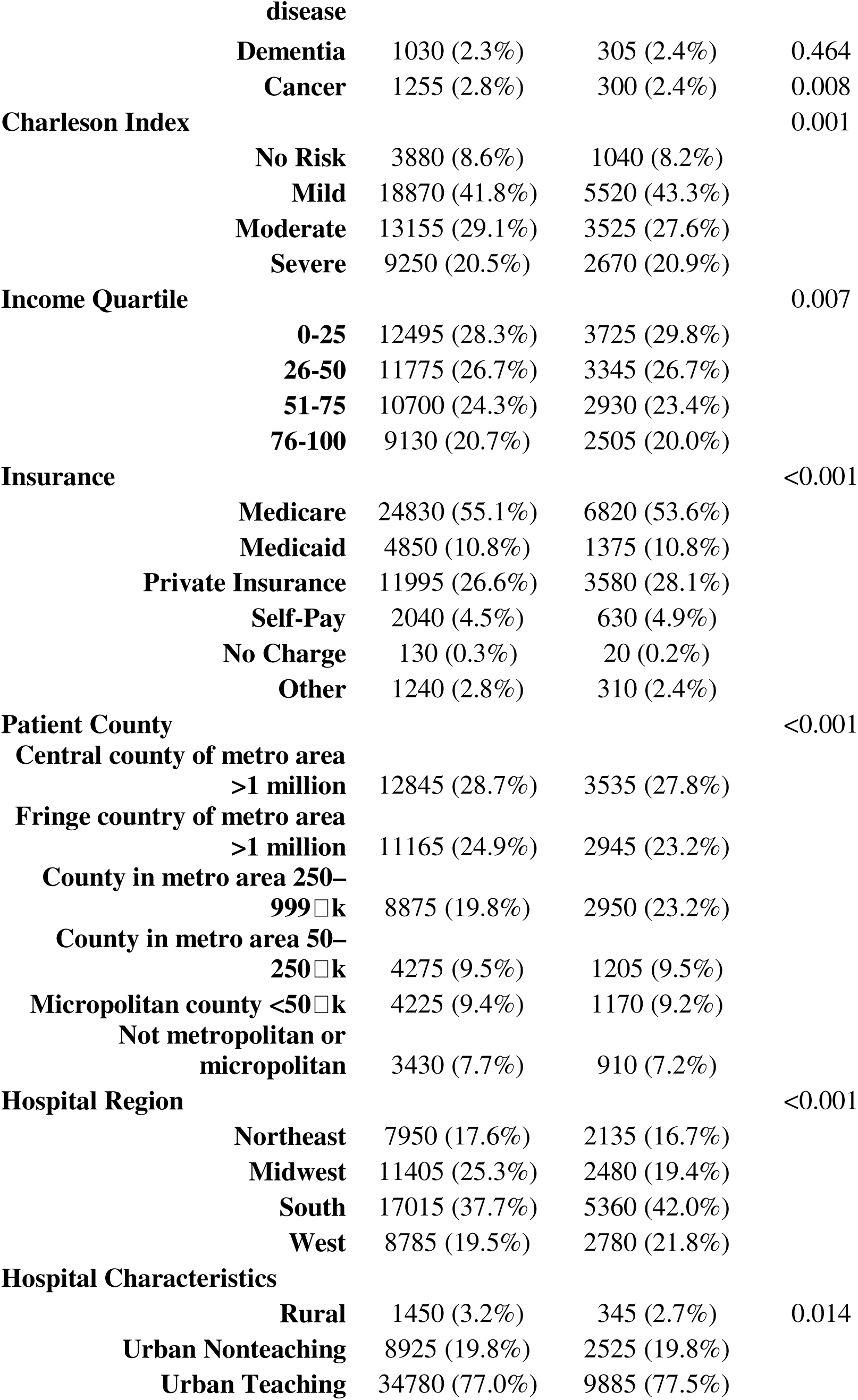
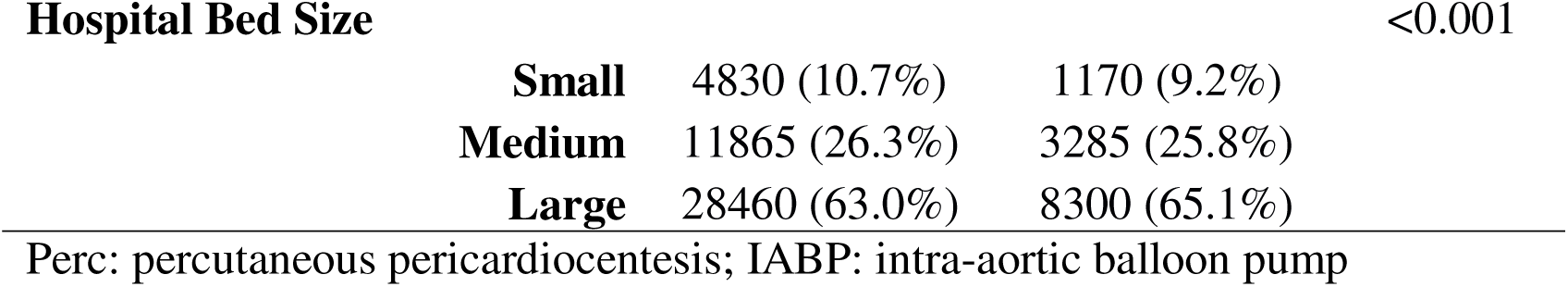
Sample patient and hospital characteristics.

Significant regional variation was observed in procedural utilization, clinical outcomes, and resource utilization. The South had the highest use of percutaneous ventricular assist devices, including Impella and TandemHeart (23.9% combined), whereas Midwest demonstrated the greatest reliance on Intra-aortic balloon pump (82.1%) (p<0.001). Length of stay was longest in the Northeast (14.5 days) and shortest in the West (11.8 days) (p<0.001). In-hospital mortality was high across all regions, ranging from 31.0% in the Midwest to 33.8% in the West (p<0.001). The West incurred the highest mean total hospital charges ($448,334), followed by the Northeast ($363,861), South ($317,249), and Midwest ($264,899) (p<0.001). Complication rates also varied regionally, with higher rates of respiratory failure in the West (64.9%), cardiac arrest in the Midwest and South (19.8% each), and dialysis utilization in the Midwest (9.0%) (all p<0.001). Discharge disposition differed significantly, with higher rates of home discharge in the South (24.2%) and West (23.3%) compared with the Northeast (15.0%) (p<0.001) (Table 2).

**Table 2:** comparison of hospital course, outcomes and disposition between regions.

| Variable | Northeast | Midwest | South | West | p-value |
| --- | --- | --- | --- | --- | --- |
| **Impella** | 355 (3.5%) | 395 (2.8%) | 770 (3.4%) | 380 (3.3%) | 0.008 |
| **TandemHeart** | 1780 (17.6%) | 2085 (15.0%) | 4590 (20.5%) | 2400 (20.8%) | <0.001 |
| **IABP** | 7950 (78.8%) | 11405 (82.1%) | 17015 (76.0%) | 8785 (76.0%) | <0.001 |
| **Length of stay (days ± SD)** | 14.5 ± 0.44 | 12.2 ± 0.23 | 12.9 ± 0.18 | 11.8 ± 0.31 | <0.001 |
| **DC Disposition** |  |  |  |  | <0.001 |
| **Home** | 1515 (15.0%) | 3030 (21.8%) | 5405 (24.2%) | 2695 (23.3%) |  |
| **SNF** | 1045 (10.4%) | 1220 (8.8%) | 2045 (9.2%) | 1245 (10.8%) |  |
| **LTAC** | 2465 (24.5%) | 3435 (24.8%) | 4685 (21.0%) | 2270 (19.6%) |  |
| **IRF** | 1785 (17.7%) | 1850 (13.3%) | 2700 (12.1%) | 1380 (11.9%) |  |
| **AMA** | 25 (0.2%) | 45 (0.3%) | 80 (0.4%) | 65 (0.6%) |  |
| **In-Hospital mortality** | 3235 (32.1%) | 4295 (31.0%) | 7410 (33.2%) | 3910 (33.8%) | <0.001 |
| **Total Hospital Charge in \$ (± SD)** | 363,861 ± 4,427 | 264,899 ± 2,392 | 317,249 ± 2,223 | 448,334 ± 5,494 | <0.001 |
| **Shock** | 8565 (84.9%) | 11560 (83.3%) | 18905 (84.5%) | 9920 (85.8%) | 0.001 |
| **Bleeding** | 2705 (26.8%) | 4005 (28.8%) | 5930 (26.5%) | 2895 (25.0%) | <0.001 |
| **AKI** | 5245 (52.0%) | 7480 (53.9%) | 11980 (53.5%) | 6135 (53.0%) | 0.025 |
| **Dialysis** | 690 (6.8%) | 1255 (9.0%) | 1675 (7.5%) | 975 (8.4%) | <0.001 |
| **Sepsis** | 1595 (15.8%) | 2020 (14.5%) | 3570 (16.0%) | 2005 (17.3%) | <0.001 |
| **Pneumonia** | 1645 (16.3%) | 2315 (16.7%) | 4220 (18.9%) | 2235 (19.3%) | <0.001 |
| **Resp Failure** | 5575 (55.3%) | 8610 (62.0%) | 13900 (62.1%) | 7505 (64.9%) | <0.001 |
| **Cardiac Arrest** | 1695 (16.8%) | 2745 (19.8%) | 4435 (19.8%) | 2165 (18.7%) | <0.001 |
| **Arrhythmia** | 6190 (61.4%) | 9020 (65.0%) | 13550 (60.6%) | 7100 (61.4%) | <0.001 |
| **DVT** | 430 (4.3%) | 715 (5.1%) | 1045 (4.7%) | 470 (4.1%) | <0.001 |
| **PE** | 150 (1.5%) | 215 (1.5%) | 350 (1.6%) | 230 (2.0%) | 0.008 |

IABP: intra-aortic balloon pump; SNF: skilled nursing facility; LTAC: long-term acute care; IRF: inpatient rehab facility; AMA: against medical advice; AKI: Acute Kidney Injury; DVT: Deep Vein Thrombosis; PE: Pulmonary Embolism; Two-way ANOVA with Standard Deviation (SD) was used to analyze total hospital charge

In adjusted analyses, patient demographics and clinical characteristics were significantly associated with adverse outcomes. Increased comorbidity burden was associated with higher odds of in-hospital mortality (OR 1.89, p<0.001), non-home discharge (OR 1.49, p<0.001), and prolonged length of stay (OR 0.83, p<0.001). Patients also demonstrated increased risk of major complications, including acute kidney injury (OR 1.31), respiratory failure (OR 1.51), cardiac arrest (OR 1.21), arrhythmia (OR 1.17), sepsis (OR 1.21), and need for dialysis (OR 1.16) (all p<0.001). In contrast, bleeding was modestly reduced (OR 0.95, p=0.022), while no significant differences were observed for shock, pneumonia, deep vein thrombosis, or pulmonary embolism. Overall, these findings demonstrate that patient-level factors remain strong determinants of both clinical outcomes and resource utilization in this critically ill cohort (Figure 2).

**Figure 2:**
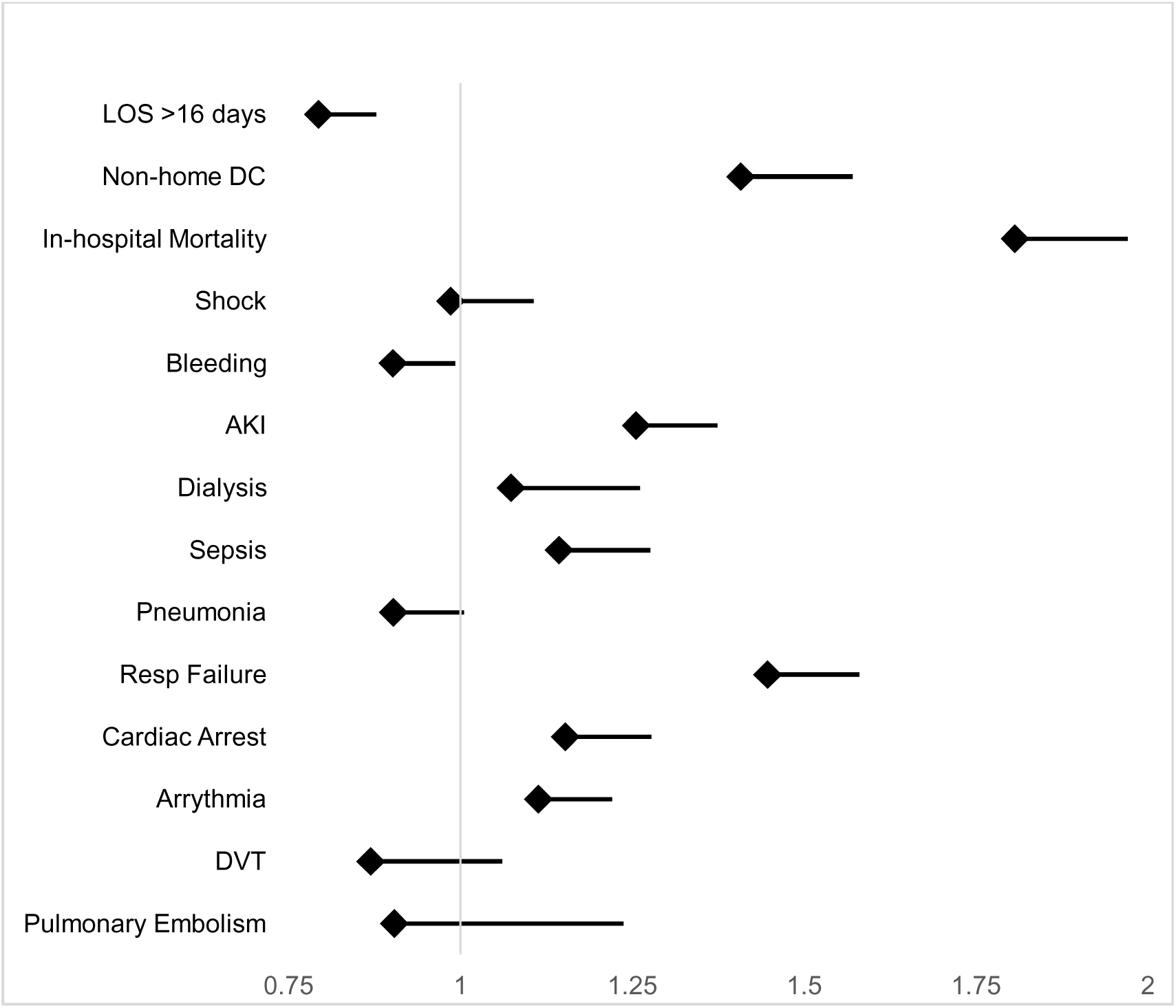
Adjusted regression of the impact of patient demographics on hospital outcomes. Adjusted for sex, race, hypertension, diabetes mellitus, valvular disease, obesity, congestive heart failure, atrial fibrillation, COPD, pulmonary hypertension, CKD, ESRD, liver disease, Charleson Comorbidity Index, income quartile, insurance status, hospital region, hospital type and hospital bed size; LOS, length of stay; DC, discharge, AKI, acute kidney injury; DVT, deep vein thrombosis. p-value ≤5 considered significant.

Mean total hospital charges varied substantially by both region and procedure type. Across all regions, percutaneous left ventricular assist device (pLVAD) use was associated with higher costs compared with Intra-aortic balloon pump. Nationally, mean charges were $403,731 for pLVAD versus $320,769 for IABP. Regional variation was pronounced, with the West demonstrating the highest costs for both pLVAD ($535,863) and IABP ($420,160), while the Midwest had the lowest costs for both procedures ($265,292 and $264,815, respectively).

Intermediate costs were observed in the Northeast and South, with consistently higher expenditures associated with pLVAD across all regions (p<0.001) (Figure 3).

**Figure 3:**
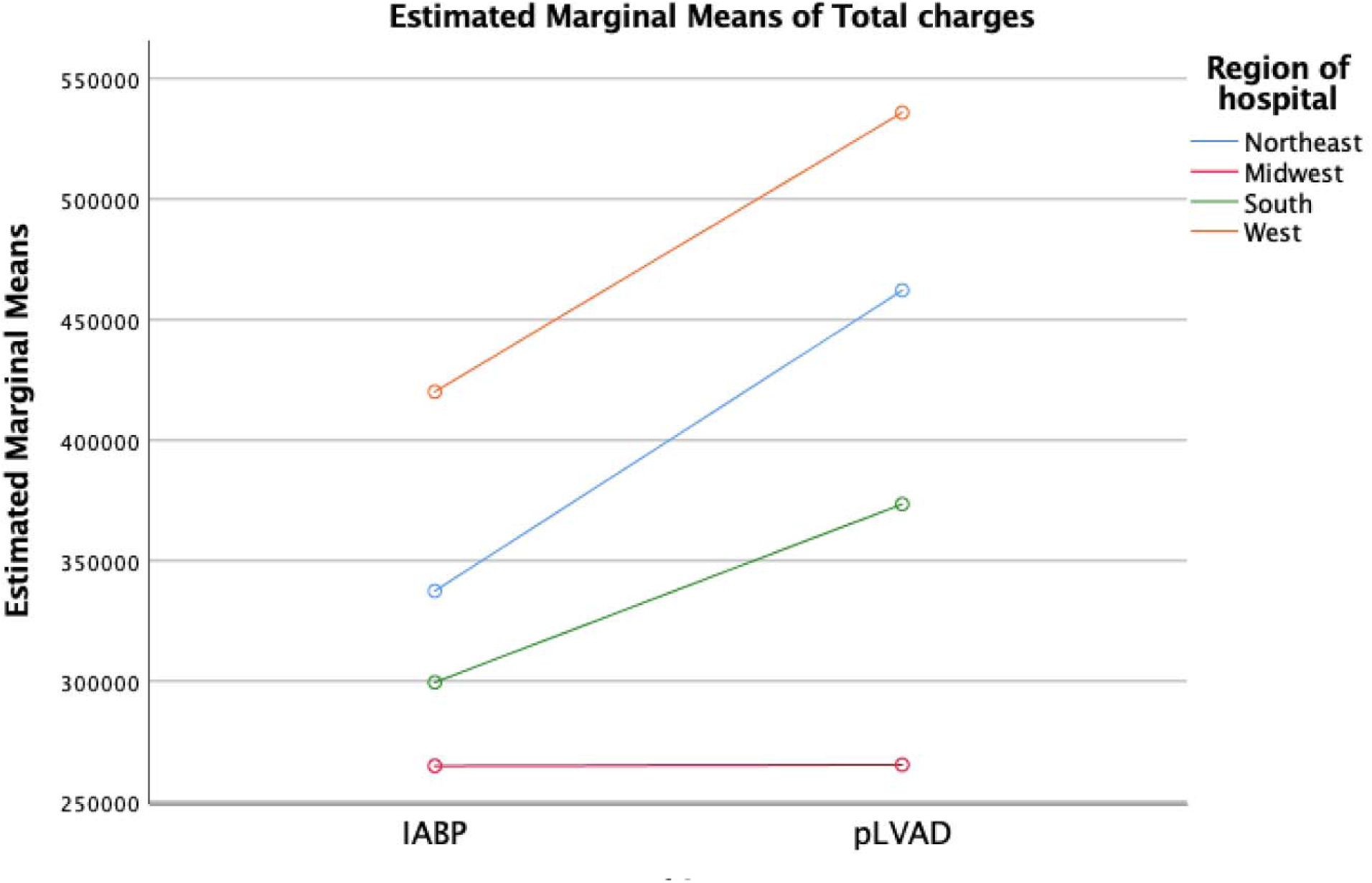
Mean total hospital charge by region for both IABP and pLVAD admissions

In adjusted analyses, multiple patient- and hospital-level characteristics were independently associated with increased hospital costs. Greater comorbidity burden was a strong driver of higher costs, including chronic kidney disease (OR 1.28), end-stage renal disease (OR 1.56), cirrhosis (OR 1.40), dementia (OR 1.25), and higher Charlson comorbidity index (severe vs none: OR 1.21; all p<0.001). Additional clinical factors associated with increased costs included prior stroke (OR 1.21), diabetes mellitus (OR 1.11), pulmonary hypertension (OR 1.15), and congestive heart failure (OR 1.04). Demographic differences were also observed, with higher costs among Black patients (OR 1.12, p<0.001) and lower costs among female patients (OR 0.89, p<0.001). Socioeconomic and payer factors influenced cost, with self-pay status associated with higher costs (OR 1.18), while Medicaid was associated with lower costs (OR 0.94) (all p<0.001). Geographic variation was significant, with higher costs observed in the Midwest (OR 1.25), South (OR 1.26), and West (OR 1.32) compared with the Northeast (all p<0.001). Additionally, non-metropolitan residence and smaller hospital settings were associated with relatively higher costs, while treatment at larger hospitals was associated with lower adjusted costs (Table 3).

**Table 3:** Impact of patient demographics on total hospital charge.

| **Variables** | **Odds Ratio**<br>**(OR)** | **95% confidence**<br>**interval** | **p-value** |
| --- | --- | --- | --- |
| Age (10-year increments) | 0.964 | 0.959–0.968 | <0.001 |
| Sex (female vs. male) | 0.891 | 0.873–0.905 | <0.001 |
| **Race (vs. White)** |  |  |  |
| Black | 1.117 | 1.079–1.126 | <0.001 |
| Asian or Pacific Islander | 0.962 | 0.930–0.988 | 0.004 |
| Hispanic | 1.02 | 0.996–1.039 | 0.078 |
| Other | 0.943 | 0.912–0.977 | <0.001 |
| Hypertension | 1.009 | 0.983–1.124 | 0.112 |
| Diabetes Mellitus | 1.108 | 1.031–1.184 | 0.003 |
| Prior Stroke | 1.212 | 1.092–1.348 | <0.001 |
| Current Smoking | 1.078 | 1.013–1.148 | 0.086 |
| Valvular Disease | 1.002 | 0.973–1.029 | 0.203 |
| Obesity | 0.989 | 0.972–1.008 | 0.074 |
| Congestive Heart Failure | 1.037 | 1.008–1.069 | 0.037 |
| Atrial Fibrillation | 0.985 | 0.967–1.005 | 0.066 |
| COPD | 1.034 | 1.011–1.056 | 0.029 |
| Pulmonary Hypertension | 1.152 | 1.102–1.201 | 0.001 |
| Chronic Kidney Disease | 1.276 | 1.237–1.329 | <0.001 |
| ESRD | 1.563 | 1.318–1.804 | <0.001 |
| Cirrhosis/Chronic liver disease | 1.396 | 1.290–1.487 | <0.001 |
| Dementia | 1.250 | 1.193–1.316 | <0.001 |
| Cancer | 1.084 | 1.037–1.145 | 0.013 |
| **Severity of illness (vs. No Risk) (CCI)** |  |  |  |
| Mild | 1.062 | 1.044–1.081 | <0.001 |
| Moderate | 1.117 | 1.082–1.153 | <0.001 |
| Severe | 1.209 | 1.117–1.289 | <0.001 |
| **Median income in patient zip code (vs. first quartile)** |  |  |  |
| Second quartile | 0.967 | 0.951–0.992 | 0.002 |
| Third quartile | 1.015 | 0.986–1.039 | 0.291 |
| Fourth quartile | 1.033 | 1.004–1.054 | 0.018 |
| **Insurance (vs. private)** |  |  |  |
| Medicare | 1.021 | 0.996–1.052 | 0.127 |
| Medicaid | 0.941 | 0.937–0.975 | <0.001 |
| Self-pay | 1.176 | 1.123–1.212 | <0.001 |
| Other | 1.068 | 1.026–1.107 | <0.001 |
| **Patient county of residence (vs. central county of metro area >1 million)** |  |  |  |
| Fringe county of metro area >1 million | 1.041 | 1.023–1.068 | <0.001 |
| County in metro area 250–999□k | 1.016 | 0.997–1.035 | 0.102 |
| County in metro area 50–250□k | 1.096 | 1.067–1.126 | <0.001 |
| Micropolitan county (<50□k) | 1.099 | 1.069–1.130 | <0.001 |
| Not metropolitan or micropolitan | 1.142 | 1.107–1.178 | <0.001 |
| **Hospital region (vs. Northeast)** |  |  |  |
| Midwest | 1.248 | 1.221–1.276 | <0.001 |
| South | 1.259 | 1.224–1.292 | <0.001 |
| West | 1.318 | 1.289–1.347 | <0.001 |
| **Hospital type (vs. rural)** |  |  |  |
| Urban non-teaching | 0.993 | 0.972–1.027 | 0.085 |
| Urban teaching | 0.984 | 0.965–0.999 | 0.041 |
| **Bed-size (vs. small)** |  |  |  |
| Medium | 0.933 | 0.912–0.952 | <0.001 |
| Large | 0.926 | 0.908–0.943 | <0.001 |

p-value ≤5 significant, determined using a gamma distribution with a log-link function.

Hospital characteristics were significantly associated with prolonged length of stay and discharge disposition. Compared with the Northeast, patients treated in the Midwest (OR 0.90, p<0.001) and West (OR 0.87, p<0.001) had lower odds of prolonged length of stay, while no significant difference was observed in the South. In contrast, all regions demonstrated significantly lower odds of non-home discharge, including the Midwest (OR 0.63), South (OR 0.55), and West (OR 0.58) (all p<0.001). Larger hospital size was strongly associated with prolonged length of stay, with medium (OR 1.29) and large hospitals (OR 2.29) demonstrating increased odds compared with small hospitals (p<0.001), while large hospitals were associated with lower odds of non-home discharge (OR 0.89, p=0.001). Compared with rural hospitals, both urban non-teaching (OR 0.29) and urban teaching hospitals (OR 0.45) were associated with significantly lower odds of prolonged length of stay (p<0.001), with no significant differences observed in discharge disposition by hospital teaching status (Table 4).

**Table 4:** Impact of hospital type on prolonged length of stay and non-home discharge.

| Hospital characteristic | Prolonged length of stay ($\geq 16$ days) | | | Non-home discharge (vs. home) | | |
| --- | --- | --- | --- | --- | --- | --- |
|  | OR | 95% CI | p-value | OR | 95% CI | p-value |
| **Region (vs. Northeast)** |  |  |  |  |  |  |
| Midwest | 0.901 | 0.850-0.955 | <0.001 | 0.632 | 0.590-0.677 | <0.001 |
| South | 1.035 | 0.982-1.091 | 0.203 | 0.550 | 0.517-0.586 | <0.001 |
| West | 0.874 | 0.822-0.930 | <0.001 | 0.581 | 0.542-0.624 | <0.001 |
| **Bed-size (vs. small)** |  |  |  |  |  |  |
| Medium | 1.288 | 1.192-1.391 | <0.001 | 0.955 | 0.886-1.028 | 0.221 |
| Large | 2.285 | 2.13-2.450 | <0.001 | 0.889 | 0.831-0.952 | 0.001 |
| **Hospital type (vs. rural)** |  |  |  |  |  |  |
| Urban non-teaching | 0.292 | 0.254-0.335 | <0.001 | 1.062 | 0.946-1.192 | 0.309 |
| Urban teaching | 0.450 | 0.427-0.475 | <0.001 | 1.032 | 0.981-1.085 | 0.224 |

Prolonged length of stay (LOS) determined as >75% percentile

In value-of-care analyses integrating risk-adjusted costs and outcomes, both Intra-aortic balloon pump and percutaneous left ventricular assist devices (pLVAD) demonstrated clinical outcomes that were better than expected after adjustment, with observed-to-expected (O/E) outcome ratios of approximately 0.92 for IABP and 0.90 for pLVAD. However, both procedures were associated with higher-than-expected costs, with O/E cost ratios exceeding 1.0 for both strategies (IABP ∼1.41 vs pLVAD ∼1.37). Despite a slightly greater relative cost inflation observed with IABP, absolute costs remained substantially lower compared with pLVAD ($210,000 vs $260,000). In value-space analysis, both strategies were positioned below the expected outcome threshold, indicating improved clinical performance; however, IABP was in the lower-cost quadrant, whereas pLVAD remained associated with higher costs without a commensurate improvement in outcomes. Collectively, these findings demonstrate that while both modalities achieve favorable risk-adjusted outcomes, IABP is associated with a more favorable overall value profile due to lower absolute cost (Figures 4-5).

**Figure 4:**
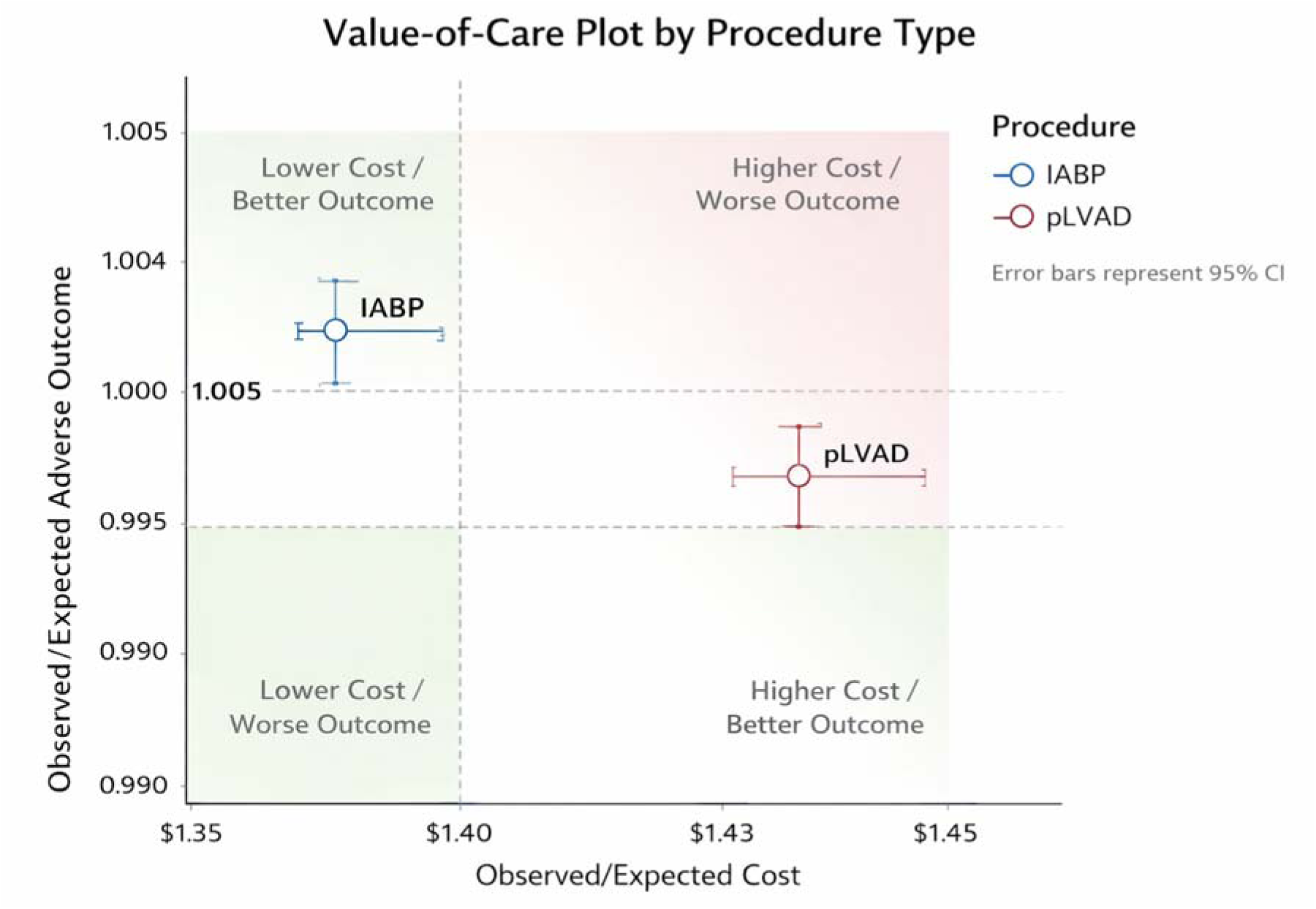
Value of care plot by procedure type

**Figure 5:**
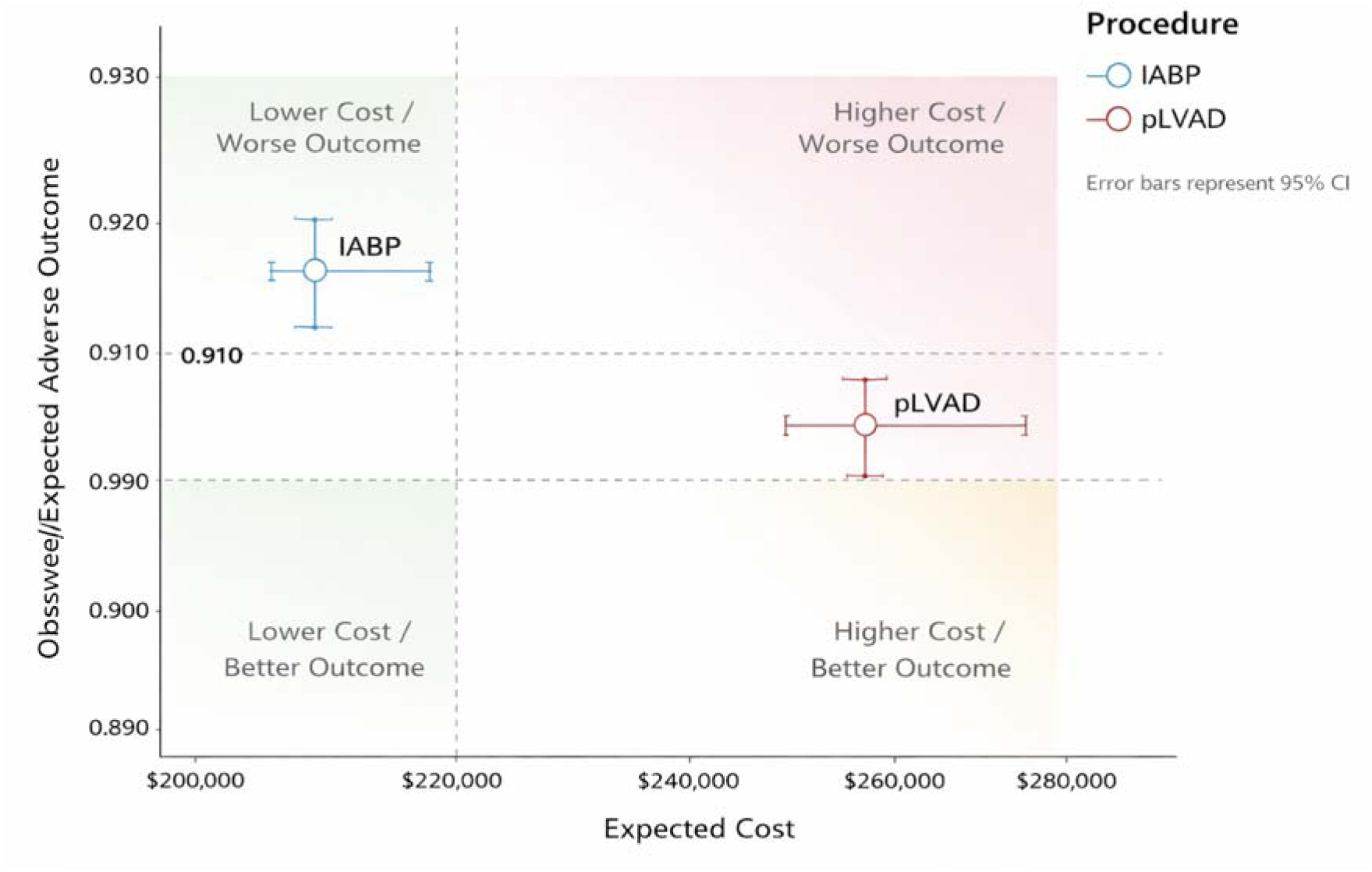
Value of expected cost compared with adverse outcome ratio

## Discussion

In this nationally representative analysis of patients undergoing percutaneous mechanical circulatory support (pMCS), we demonstrate substantial variation in resource utilization, clinical outcomes, and overall value across device strategies and regions. Leveraging contemporary ICD-10 coding within the National Inpatient Sample, this study provides the first large-scale evaluation integrating risk-adjusted costs and outcomes into a unified value-of-care framework. The principal findings are twofold: first, significant geographic and institutional variability exists in both cost and outcomes; and second, more advanced pMCS strategies are associated with higher costs without proportional improvements in clinical outcomes.

Our cohort reflects a critically ill population with high comorbidity burden and substantial mortality, exceeding 30% across all regions. These findings are consistent with prior studies of cardiogenic shock and hemodynamically unstable patients, where outcomes remain poor despite advances in mechanical support (7, 8). Although baseline characteristics between intra-aortic balloon pump (IABP) and percutaneous left ventricular assist device (pLVAD) cohorts were broadly similar, modest differences, such as higher rates of congestive heart failure in the pLVAD group, likely reflect differences in disease severity and clinical selection. The predominance of care delivered in urban teaching and large hospitals further highlights the concentration of advanced cardiac care in high-resource centers (9–11).

We observed marked regional variation in procedural utilization, outcomes, and cost. The South and West demonstrated greater utilization of pLVAD strategies, whereas the Midwest relied more heavily on Intra-aortic balloon pump. Notably, the West was associated with the highest hospital charges, while the Midwest consistently demonstrated the lowest costs. These findings align with prior literature across multiple procedural domains demonstrating higher healthcare expenditures in Western regions and lower costs in the Midwest (11, 12). Potential explanations include regional differences in labor costs, hospital market dynamics, regulatory environments, and resource availability, although these factors are not fully captured in administrative datasets (13).

Beyond geographic variation, both patient- and hospital-level characteristics were important determinants of cost and outcomes. Increased comorbidity burden, particularly renal disease, liver disease, and greater illness severity, was strongly associated with higher costs, consistent with prior studies in critically ill populations (10, 11). Additionally, smaller hospitals and non-metropolitan settings were associated with relatively higher adjusted costs, potentially reflecting reduced economies of scale and limited access to specialized infrastructure. Larger hospitals, in contrast, demonstrated relatively lower costs, suggesting potential efficiency gains associated with higher procedural volume and resource availability (14).

The most clinically and policy-relevant finding of this study is the divergence in value between pMCS strategies. Although both IABP and pLVAD were associated with clinical outcomes that were comparable to or better than expected after risk adjustment, pLVAD use was associated with substantially higher costs without a proportional reduction in adverse outcomes. In contrast, IABP achieved similar or slightly better-than-expected outcomes at significantly lower cost, resulting in a more favorable value profile. These findings are consistent with prior clinical trials and observational studies that have questioned the incremental benefit of more advanced mechanical support devices despite increased resource utilization (8, 9, 15). Importantly, our results extend this literature by quantifying this discrepancy within a national, risk-adjusted, value-based framework.

This study contributes to the evolving emphasis on value-based care in cardiovascular medicine by introducing a novel approach that integrates observed-to-expected cost and outcome metrics. While prior investigations have largely evaluated outcomes or costs in isolation, our framework provides a more comprehensive assessment of procedural efficiency and may better inform clinical decision-making, hospital resource allocation, and policy development (16, 17). As healthcare systems increasingly prioritize value, such approaches may be critical in guiding appropriate utilization of high-cost technologies.

A critical consideration in interpreting these findings is the potential for residual confounding due to differences in illness severity and clinical indication. Patients receiving percutaneous left ventricular assist devices (pLVAD) are often more critically ill, with more advanced cardiogenic shock, failure of initial therapies, or need for escalation of support, factors that are not fully captured in administrative datasets such as the National Inpatient Sample (9, 18). Important clinical variables, including hemodynamic parameters, Society for Cardiovascular Angiography and Interventions (SCAI) shock stage, timing of device initiation, and use of stepwise escalation strategies, are not available and may significantly influence both outcomes and costs (19). As such, the observed differences in cost and outcomes between intra-aortic balloon pump (IABP) and pLVAD should not be interpreted as causal or reflective of intrinsic device effectiveness, but rather as population-level associations within a heterogeneous and high-risk cohort.

These findings should also be interpreted within the context of evolving management paradigms in cardiogenic shock. Contemporary practice increasingly emphasizes early identification, multidisciplinary “shock team” approaches, and tailored escalation of mechanical circulatory support based on patient-specific factors (9, 20). In this framework, pLVAD use may be clinically appropriate in select patients despite higher costs, particularly when used for more severe or refractory shock (21). The higher utilization of pMCS at large academic and tertiary referral centers likely reflects referral patterns and case complexity, as these institutions frequently manage patients with more advanced cardiogenic shock and greater clinical acuity.

Notably, both IABP and pLVAD cohorts in this study demonstrated outcomes that were better than expected after risk adjustment, suggesting that advances in systems of care and patient selection may be contributing to improved survival in this population. However, the absence of a proportional improvement in outcomes relative to cost at the population level highlights the need to better define which patients derive the greatest incremental benefit from advanced support strategies.

Mechanistically, the higher costs observed with pLVAD are likely multifactorial, reflecting not only the intrinsic cost of the device itself but also increased procedural complexity, longer intensive care utilization, and higher rates of complications such as acute kidney injury, bleeding, and respiratory failure. Additionally, institutional factors, including operator experience, device availability, and hospital-level practice patterns, may contribute to variability in both utilization and efficiency (9). These findings underscore the importance of optimizing patient selection and standardizing care pathways to ensure that advanced mechanical support is deployed in a manner that maximizes clinical benefit while minimizing unnecessary resource utilization (19, 22). Future studies incorporating granular clinical data and prospective designs will be essential to more precisely define the role of pMCS strategies within a value-based framework.

Several limitations warrant consideration. As a retrospective analysis of administrative data from the National Inpatient Sample, this study is subject to coding inaccuracies and lacks granular clinical detail, including hemodynamics, laboratory values, timing of intervention, and device-specific indications, introducing potential residual confounding and selection bias. Hospital charges were used as a surrogate for true costs and may not accurately reflect resource utilization, and the NIS does not provide cost breakdowns or capture prehospital and post-discharge care, limiting comprehensive cost assessment. The dataset is restricted to inpatient hospitalizations, which may overrepresent critically ill patients and influence measures such as length of stay and admission type. Additionally, variation in coding practices, institutional protocols, operator experience, and device availability may contribute to heterogeneity in both cost and outcomes. The lack of longitudinal follow-up precludes assessment of long-term outcomes and cost-effectiveness. Finally, the use of early ICD-10 era data may not fully reflect contemporary practice patterns or stabilized cost structures. These findings should therefore be interpreted as hypothesis-generating.

## Conclusion

We demonstrate significant variation exists in the cost, outcomes, and overall value of pMCS strategies across the United States. While pLVAD use is associated with higher costs, it does not confer proportional improvements in clinical outcomes compared with IABP, raising important considerations regarding optimal resource utilization in critically ill patients. Future studies should focus on refining patient selection and incorporating value-based frameworks to optimize both clinical outcomes and healthcare efficiency.

## Data Availability

Data available on reasonable request.

## Ethics Statement

Ethical approval was not sought for this study as it utilized exclusively pre-existing, de-identified, and publicly available data.

## Conflicts of Interests

None

## Author Contributions

Conceptualization: JG, WA, JL, ZT

Methodology: JG, WA, JL, ZT

Analysis: JG, AH

Manuscript writing: JG, WA, AH

Critical Revisions: JG, AH

## Funding

None

## Data Availability

All data produced in the present study are available upon reasonable request to the authors

## Notes

### Competing Interest Statement

The authors have declared no competing interest.

## References

1. Tsao CW, Aday AW, Almarzooq ZI, et al. Heart Disease and Stroke Statistics-2023 Update: A Report From the American Heart Association. Circulation. 2023; 147: e93–e621.

2. Himmelstein DU, Thorne D, Warren E, et al. Medical bankruptcy in the United States, 2007: results of a national study. Am J Med. 2009; 122: 741–6.

3. Papanicolas I, Woskie LR, Jha AK. Health Care Spending in the United States and Other High-Income Countries. Jama. 2018; 319: 1024–39.

4. Kannan S, Stevens J, Song Z. Growth In Patient Cost Sharing For Hospitalizations With And Without Intensive Care Among Commercially Insured Patients. Health Aff (Millwood). 2023; 42: 1221–29.

5. Malik A, Basu T, VanAken G, et al. National Trends for Temporary Mechanical Circulatory Support Utilization in Patients With Cardiogenic Shock From Decompensated Chronic Heart Failure: Incidence, Predictors, Outcomes, and Cost. J Soc Cardiovasc Angiogr Interv. 2023; 2: 101177.

6. Gilani FS, Farooqui S, Doddamani R, et al. Percutaneous Mechanical Support in Cardiogenic Shock: A Review. Clin Med Insights Cardiol. 2015; 9: 23–8.

7. Thiele H, Zeymer U, Thelemann N, et al. Intraaortic Balloon Pump in Cardiogenic Shock Complicating Acute Myocardial Infarction: Long-Term 6-Year Outcome of the Randomized IABP-SHOCK II Trial. Circulation. 2019; 139: 395–403.

8. Schrage B, Ibrahim K, Loehn T, et al. Impella Support for Acute Myocardial Infarction Complicated by Cardiogenic Shock. Circulation. 2019; 139: 1249–58.

9. Henry TD, Tomey MI, Tamis-Holland JE, et al. Invasive Management of Acute Myocardial Infarction Complicated by Cardiogenic Shock: A Scientific Statement From the American Heart Association. Circulation. 2021; 143: e815–e29.

10. Vallabhajosyula S, Dunlay SM, Prasad A, et al. Acute Noncardiac Organ Failure in Acute Myocardial Infarction With Cardiogenic Shock. J Am Coll Cardiol. 2019; 73: 1781–91.

11. Khera R, Cram P, Vaughan-Sarrazin M, et al. Use of Mechanical Circulatory Support in Percutaneous Coronary Intervention in the United States. Am J Cardiol. 2016; 117: 10–6.

12. Dieleman JL, Cao J, Chapin A, et al. US Health Care Spending by Payer and Health Condition, 1996-2016. Jama. 2020; 323: 863–84.

13. Cooper Z, Craig SV, Gaynor M, et al. THE PRICE AIN’T RIGHT? HOSPITAL PRICES AND HEALTH SPENDING ON THE PRIVATELY INSURED. Q J Econ. 2019; 134: 51–107.

14. Finks JF, Osborne NH, Birkmeyer JD. Trends in hospital volume and operative mortality for high-risk surgery. N Engl J Med. 2011; 364: 2128–37.

15. Ouweneel DM, Eriksen E, Sjauw KD, et al. Percutaneous Mechanical Circulatory Support Versus Intra-Aortic Balloon Pump in Cardiogenic Shock After Acute Myocardial Infarction. J Am Coll Cardiol. 2017; 69: 278–87.

16. Porter ME. What is value in health care? N Engl J Med. 2010; 363: 2477–81.

17. Herzlinger RE. How to solve the cost crisis in health care. Harv Bus Rev. 2011; 89: 22, discussion 23.

18. van Diepen S, Katz JN, Albert NM, et al. Contemporary Management of Cardiogenic Shock: A Scientific Statement From the American Heart Association. Circulation. 2017; 136: e232–e68.

19. Naidu SS, Baran DA, Jentzer JC, et al. SCAI SHOCK Stage Classification Expert Consensus Update: A Review and Incorporation of Validation Studies: This statement was endorsed by the American College of Cardiology (ACC), American College of Emergency Physicians (ACEP), American Heart Association (AHA), European Society of Cardiology (ESC) Association for Acute Cardiovascular Care (ACVC), International Society for Heart and Lung Transplantation (ISHLT), Society of Critical Care Medicine (SCCM), and Society of Thoracic Surgeons (STS) in December 2021. J Soc Cardiovasc Angiogr Interv. 2022; 1: 100008.

20. Taleb I, Koliopoulou AG, Tandar A, et al. Shock Team Approach in Refractory Cardiogenic Shock Requiring Short-Term Mechanical Circulatory Support: A Proof of Concept. Circulation. 2019; 140: 98–100.

21. Tongers J, Sieweke JT, Kühn C, et al. Early Escalation of Mechanical Circulatory Support Stabilizes and Potentially Rescues Patients in Refractory Cardiogenic Shock. Circ Heart Fail. 2020; 13: e005853.

22. Tehrani BN, Truesdell AG, Psotka MA, et al. A Standardized and Comprehensive Approach to the Management of Cardiogenic Shock. JACC Heart Fail. 2020; 8: 879–91.

